# Circadian instability predicts PTSD symptom severity following mass trauma

**DOI:** 10.1101/2025.03.19.25324240

**Authors:** Noa Magal, Ophir Netzer, Eden Eldar, Noga Mandelblit, Nitsan Sagi, Michal Oren, Roy Salomon, Roee Admon

## Abstract

**Objective:** Trauma exposure may lead to posttraumatic stress disorder (PTSD) in a subset of vulnerable individuals. Circadian rhythm disruptions have emerged as both risk factors and consequences of PTSD. Whether circadian disruptions in trauma aftermath predict future PTSD risk, and if so, what behavioral and physiological indices of circadian disruption underlie risk prediction, is not yet clear.

**Methods:** Survivors of the Supernova music festival mass trauma event (*n*=211) were monitored in real-life settings for a full month, three-to-six months post-event, using wearable sensors that tracked their heart rate (HR), activity and sleep. PTSD symptoms were assessed using the Posttraumatic Checklist for DSM-5 (PCL-5) before and after the recording month, as well as at follow-up eight-to-eleven months post-event. Matched controls (*n*=113) underwent an identical procedure. Behavioral and physiological indices of circadian rhythm disruptions were quantified using interday sleep stability and cosinor HR analysis, respectively. Linear regressions tested associations between circadian measures and PTSD status and severity at follow-up.

**Results:** Compared to controls, trauma survivors exhibited circadian instability three-to-six months post-event, expressed behaviorally as reduced interday sleep stability and physiologically as higher variance in daily HR phases. Critically, 54% of trauma survivors exceeded the clinical threshold for PTSD at follow-up. Circadian instability three-to-six months post-event was particularly potent among survivors with PTSD at follow-up and predicted their symptom severity.

**Conclusions:** Specific behavioral and physiological indices of circadian instability three-to-six months post-trauma predict future PTSD risk. Maintaining stable circadian rhythms in trauma aftermath may mitigate PTSD symptoms, highlighting avenues for monitoring and early interventions.

## Introduction

Exposure to extreme trauma exerts unparalleled and profound impact across multiple aspects of daily life. For a subset of vulnerable individuals, trauma exposure may result in chronic stress-related psychopathology, typically posttraumatic stress disorder (PTSD), yielding significant suffering and disability (1). These devastating effects are particularly common following interpersonal violent events such as terror attacks. Following exposure to terror attacks, up to 39% of individuals may develop PTSD in the first six months post-event, with 22% of individuals continuing to exhibit clinical PTSD status one-year post-event (2, 3). Characterizing the mechanisms that underlie vulnerability to develop psychopathology following trauma exposure may carry substantial clinical and societal implications. For example, identifying factors in the aftermath of trauma that predict future PTSD risk may enable directing early therapeutic interventions toward vulnerable individuals (4).

Circadian rhythms, derived from Latin to mean “about a day,” are intrinsic 24- hour cycles that govern numerous processes in the human body, including heart rate (HR), blood pressure, body temperature, and hormonal release (5–8). These physiological circadian oscillations, in turn, regulate fundamental daily behaviors such as sleep-wake cycles, activity, and food intake, ensuring their occurrence at optimal times of the day. Extensive research established the robust associations between circadian rhythm disruptions and mental health disorders (9, 10). In recent years, posttraumatic chronodisruption, disturbance of the circadian rhythm following trauma exposure, has emerged as a robust pathway for trauma vulnerability (11, 12). In support of that, behavioral and physiological indices of circadian dysregulation including sleep disturbances (13) and disrupted circadian HR dynamics (14), have been reported in PTSD patients. Critically however, circadian disruptions were found to be associated with PTSD both as vulnerability markers that, if predate trauma exposure, increase the risk for future PTSD, as well as maladaptive consequences that emerge following trauma exposure (15–19). Hence, the contribution of circadian disruption to PTSD prediction is not yet clear. In addition, behavioral circadian sleep measures were shown to vary substantially in the general population in real-life environment, more within-person than between persons (19–21), with PTSD patients not differing from healthy controls in objective mean measures of sleep onset and duration (22). Therefore, mean circadian patterns may carry limited merit for vulnerability prediction. Instead, it has been suggested that indices of intra-individual variability that represent stability or lack thereof of circadian patterns over time may better indicate vulnerability (19, 23, 24).

A tragic and unique opportunity to assess the potential contribution of circadian instability to PTSD symptom severity in ecological settings has emerged following the terror attack on Israel in October 7^th^, 2023 (25). On that day, over 360 of the ∼4,000 Supernova music festival attendees were killed, and dozens were taken hostage. While fleeing the festival grounds, survivors witnessed severe, life-threatening trauma for an extended period. The combination of trauma severity, the number of individuals directly exposed to atrocities and the unique characteristic of the event is unprecedented. Initial reports indicate that more than half of individuals exposed to this mass trauma exhibited PTSD symptom severity above the clinical cutoff for PTSD three-to-six months following the event (26). The goal of the current study was to assess whether behavioral and physiological measures of circadian disruption, and particularly circadian instability, can predict long term risk for PTSD within this unique cohort. For that, 211 survivors of the mass trauma at the Supernova music festival were monitored using wearable sensors for one full month while maintaining their daily routine in real-life settings. Measurements took place three-to-six months following the event, with PTSD symptom severity assessed using a validated scale at the start and end of the measurement period, as well as at follow-up eight-to-eleven months post-event. Matched controls (*n*=113) underwent an identical procedure. Behavioral and physiological indices of circadian disruptions were quantified via sleep interday stability (IS) index and cosinor HR patterns, respectively. Given the importance of circadian consistency to mental health and coping, we reasoned that higher intra-individual interday variability in behavioral and physiological circadian indices (i.e., circadian instability) would predict PTSD status at follow-up and will be associated with more severe PTSD symptoms.

## Methods

### Participants

Out of 750 survivors that were approached by our study staff, 211 adults who survived the October 7th, 2023, Supernova music festival mass trauma event were recruited to the study, alongside 113 matched control participants. See **Table 1** for demographic information and statistical comparisons between groups. Survivors were recruited through various channels, including partnership with the non-profit support organization (NGO) “Safeheart”, referrals from friends who experienced the attack together, and survivor support groups on social media. Control group participants were recruited through social media groups that shared similar interests as the survivors (e.g. music festivals) and using referrals from friends. Eligibility criteria were liberal to allow maximum inclusion of the population investigated, with the limitations of excluding individuals who are during active military service, pregnant, and aged below 18 or above 55. Inclusion criteria for the survivors\control groups required participants to have\not have attended the Supernova music festival during the attack. Recruitment took place in various locations across Israel and included meetings with participants in designated spaces as well as home visits for those unable or unwilling to arrive. Given the sensitivity of the population being studied, special attention was paid to obtaining informed consent. First, research assistants were instructed not to pressure participants into taking part and to emphasize that participation was entirely voluntary. Participants were free to choose which aspects of the study to engage in (e.g. participants could choose to answer questionnaires without wearing the sensor and were compensated accordingly). Second, research assistants were instructed to prioritize participants’ well-being. If a participant expressed or was suspected of experiencing distress, they were referred to SafeHeart NGO or other organizations that provide treatment. All procedures adhered to the University of Haifa’s institutional guidelines and were approved by the University of Haifa Institutional Ethics Committee (approval no. 374/23).

**Table 1.** Demographics and PTSD symptoms across groups.

|  | Survivors<br>PTSD + | Survivors<br>PTSD - | Control | Survivors<br>Dropout from<br>Follow-Up | p-<br>value<sup>2,3</sup> |
| --- | --- | --- | --- | --- | --- |
| Total n | 97 | 82 | 113 | 33 |  |
| Age, mean (sd) | 26.7 (7.1)<sup>a</sup> | 28.8 (7.8) | 29.6 (7.3)<sup>a,b</sup> | 25.8 (4.4)<sup>b</sup> | **.007** |
| Female sex, n (%) | 60 (62%)<sup>a</sup> | 35 (42%) | 66 (58%) | 10 (30%)<sup>a</sup> | **.003** |
| Sensor data days, mean (sd)<sup>4</sup> | 20.5 (9.1)<sup>a,b,c</sup> | 24 (7.9)<sup>a,d,e</sup> | 28.5 (7.6)<sup>b,d,f</sup> | 14.4 (4.8)<sup>c,e,f</sup> | **<.001** |
| Sleep time minutes, mean (sd) | 477 (58.9) | 463 (49.5) | 461 (37.2) | 474 (68.4) | .122 |
| **First assessment before sensor recordings** |  |  |  |  |  |
| PCL score, mean (sd) | 48.1 (11.3)<sup>a,b,c</sup> | 26.5 (15.7)<sup>a,d</sup> | 26.9 (15.9)<sup>b,e</sup> | 40.3 (14.7)<sup>c,d,e</sup> | **<.001** |
| PTSD status, n (%) | 88 (92%)<sup>a,b</sup> | 26 (31%)<sup>a,c</sup> | 44 (40%)<sup>b,d</sup> | 24 (72%)<sup>c,d</sup> | **<.001** |
| **Second assessment after sensor recording** |  |  |  |  |  |
| PCL score, mean (sd) | 45.8 (12.9)<sup>a,b,c</sup> | 22.3 (10.4)<sup>a,d</sup> | 19.5 (14.6)<sup>b,e</sup> | 37 (15.9)<sup>c,d,e</sup> | **<.001** |
| PTSD status, n (%) | 82 (87%)<sup>a,b</sup> | 12 (14%)<sup>a,c</sup> | 27 (25%)<sup>b,d</sup> | 16 (64%)<sup>c,d</sup> | **<.001** |
| **Follow-up assessment** |  |  |  |  |  |
| PCL score, mean (sd) | 48.9 (9.7)<sup>a</sup> | 20.4 (7.9)<sup>a</sup> | - | - | **<.001** |
| PTSD status, n (%) | 97 (100%) | 0 | - | - | **<.001** |
| Weeks since event, mean (sd) | 40.7 (5.0) | 39.1 (4.4) | - | - | .057 |

<sup>1</sup>Groups sharing the same superscript letter (e.g., <sup>a</sup>) are significantly different from each other.
<sup>2</sup>For continuous variables, p-values are based on F-test (ANOVA) and comparisons are adjusted using Tukey's HSD.
<sup>3</sup>For categorical variables, p-values are based on chi square test and comparisons are adjusted using Bonferroni corrections for multiple comparisons and the fisher exact test per comparison.
<sup>4</sup>Total days with data are based on the maximum value of days in either cosinor HR model or IS calculations.

### Procedure

Following recruitment, participants completed surveys assessing demographic variables including sex and age, physical health status, current consumption of medications and additional substances and current or past clinical treatment. Participants also completed the Adverse Childhood Experience Questionnaire [ACE; (27)], and the Posttraumatic Checklist for DSM-5 [PCL-5; (28)] to assess PTSD status and symptom severity. Next, participants were provided with a wearable sensor (Fitbit Charge5) and instructed to conduct their daily routine activities without removing the sensor, including during sleep and showers, for one full month. Fitbit Charge 5 was specifically chosen for this study due to its long battery life and established reliability and validity in measuring various health aspects including sleep, activity and HR (29, 30). During the recording month participants received notifications every five days via a dedicated smartphone application reminding them to charge the sensor and synchronize it with the Fitbit cloud. If data was not synchronized, a research team member contacted the participant by phone to remind them to do so. Following the one-month recording period participants returned the sensors and completed the PCL-5 for the second time. The wearable recording period for the survivors group took place between November 21, 2023, to March 14, 2024, covering a period of three-to-six months since the attack, an aftermath period known to be critical for PTSD development and chronification or attenuation (1, 2). Then, a follow-up assessment of PCL-5 was conducted eight-to-eleven-month post event. For the control group, data collection period was extended to August 25, 2024, with majority of control group participants (77%) sampled in the period between May 2024 and August 2024.

### Data preprocessing

Fitbit data were collected using the Fitbit cloud. HR signal was received at a resolution of 5-15 seconds. Sleep data were provided in epochs indicating sleep onset, offset, and waking times during sleep. Preprocessing was performed using in-house code in Python and R programming language environment, following our previously established methods (31, 32). For HR data, preprocessing included removing samples with a low confidence score as defined by Fitbit algorithm as well as samples with beats per minute (BPM) values lower than 40 or higher than 180, and down sampling to a 5-minute resolution. For sleep data, preprocessing involved identifying sleep and wake periods and down sampling to a 5-minute resolution using a majority-based mode approach. Days with more than 37% (540 minutes) of missing data were discarded. For more information on preprocessing steps see **Supplementary material**.

### Circadian sleep assessment

Sleep was quantified using the interday stability (IS) index. IS, typically computed using activity data as sampled by actigraph, quantifies the stability of rest–activity rhythms by comparing the average activity pattern in each day per participant to their overall average pattern, and is considered more sensitive to sleep fragmentation compared to standard deviation metrics of specific sleep parameter (24). As suggested before, IS was calculated by assigning binary state (i.e. sleep = 0 and wake = 1) to each epoch (5-minue) and measuring the ratio of the variance within the same time interval in each day relative to the overall variance (24). Hence, total IS scores can range between 0 to 1, with lower scores indicating reduced sleep interday stability. **Figure 1A** depicts examples of two representative participants demonstrating high (left) vs. low (right) sleep interday stability (IS). For more information see **Supplementary material**.

**Figure 1.**
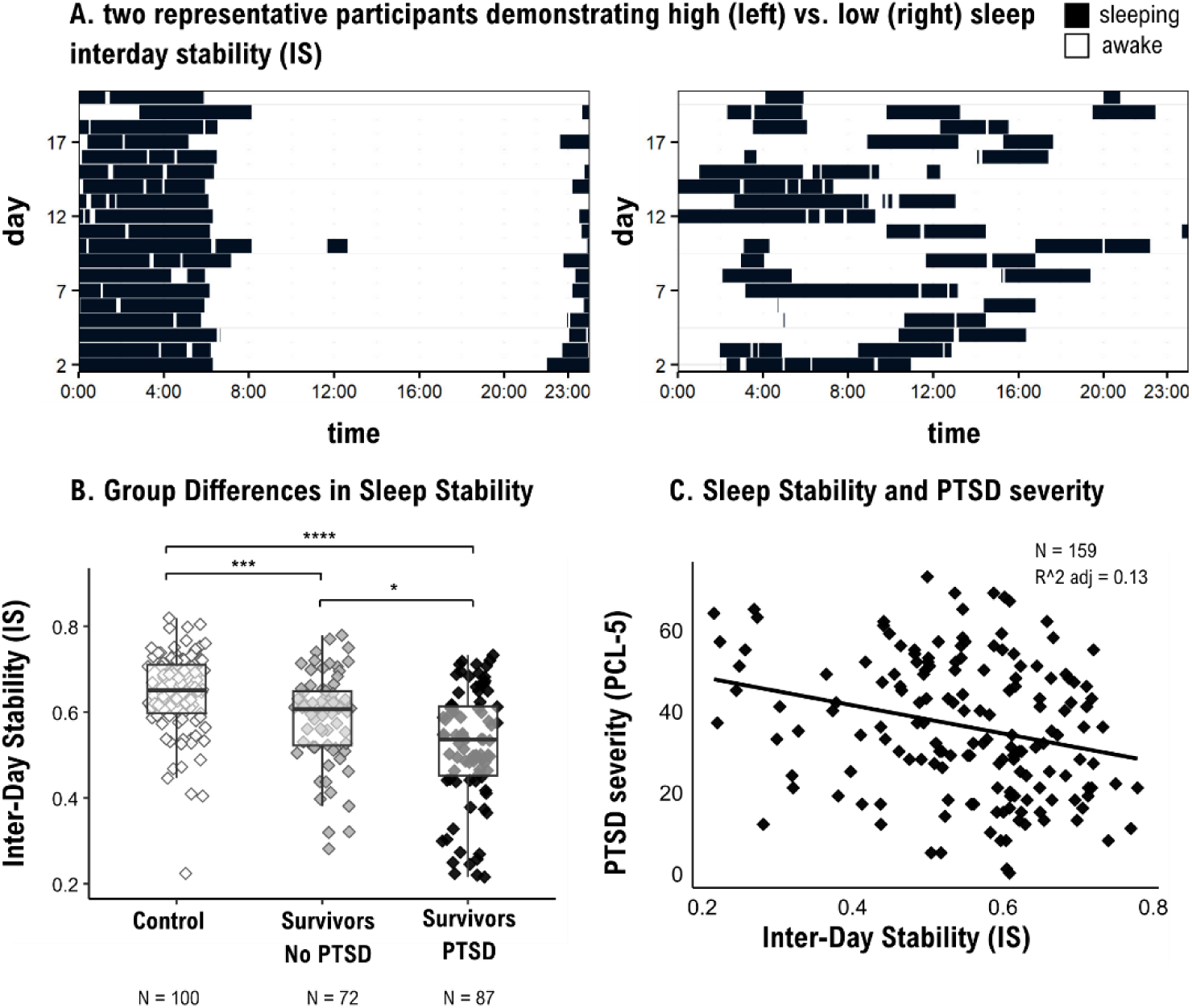
PTSD and sleep interday stability. **A)** 24h actograms from two representative participants, showing high (left) vs. low (right) sleep interday stability (IS = 0.76 vs. IS = 0.14, respectively). Black tiles are minutes labelled as “sleep” and white tiles are minutes labelled as “awake”. **B)** Linear regression model for group differences in IS uncovered lower IS values for survivors with PTSD at follow-up compared to survivors with no PTSD at follow-up as well as compared to matched controls. **C)** Linear regression model for PTSD severity and IS for the survivors group uncovered that lower IS is associated with more severe PTSD symptom severity at follow-up. *R^2 adjusted*: the adjusted R squared of the whole model. Slope represents the simple OLS slope. *IS*: interday stability of sleep. *PCL-5*: PTSD checklist for DSM-5.

### Circadian HR assessment

Circadian HR was calculated by fitting a single cosine curve to the HR BPM data of each day using a least square regression, as implanted in the ‘cosinor’ and ‘cosinor2’ packages in r (33, 34) (**Figure 2A**). To increase model accuracy, activity (steps) was included as an additional linear regressor (35). By fitting the model to each day separately, both overall means as well as standard deviations across the entire measurement period were calculated for the circadian indices of MESOR (intercept), amplitude and acrophase per participant. Standard deviation measures were log-transformed to reduce skewness before modeling. For more information see **Supplementary material**.

**Fig 2.**
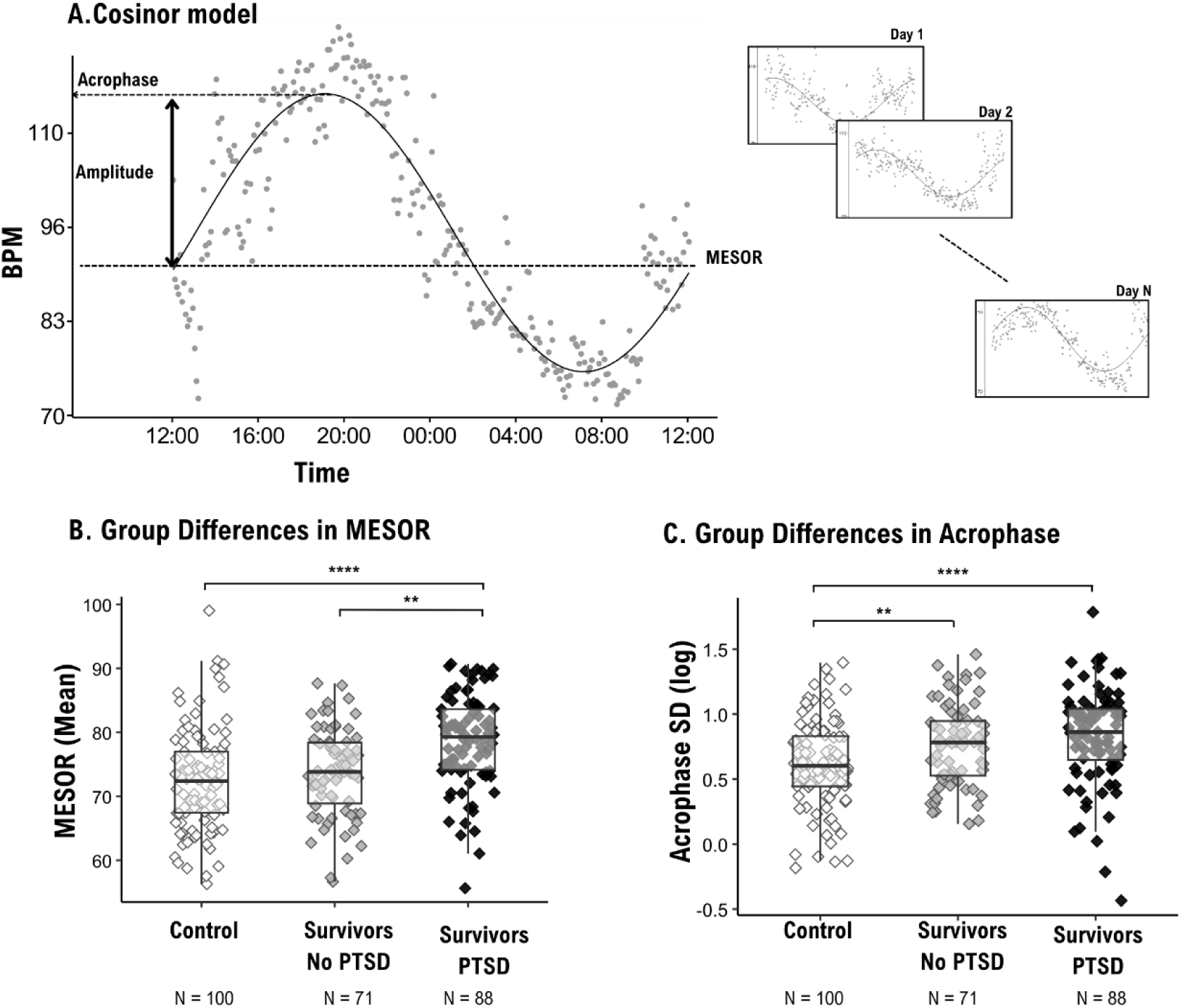
PTSD and Circadian HR. **A)** On the left is an example of a 24h heart rate data (grey dots) and the fitted cosinor model (black curve). Right: the cosine curve was repeatedly fitted to every 24h, allowing the calculation of both mean and stability estimates of circadian rhythms per participant over a full month of continuous recording in real-life settings. **B)** Linear regression model for group differences in mean MESOR uncovered higher MESOR for survivors with PTSD at follow-up compared to survivors with no PTSD at follow-up as well as compared to matched controls. **C)** Linear regression model for group differences in Acrophase SD uncovered higher values (i.e., more instability) in survivors with PTSD as well as survivors with no PTSD at follow-up compared to matched controls. *BPM:* Beats per minute; *MESOR:* Midline estimation of rhythm; *Acrophase SD:* Circular standard deviation of heart rate peak time (log transformed).

### Analytical approach

Out of the 211 survivors that were recruited to the study, 206 survivors provided wearable sensor data, with 28 of them excluded due to insufficient number of data collection days or poor data quality. Of the 113 matched controls that were recruited, 105 provided wearable sensor data, from which 6 were excluded due to insufficient number of data collection days or poor data quality. Hence, the final sample size was composed of 178 survivors and 99 control participants. Also out of the 211 survivors, 179 (85%) provided PCL-5 scores at follow-up. Survivors were divided into two groups based on whether their PCL-5 scores at follow-up, nine-to-eleven-month following the attack, were above (PTSD+) or below (PTSD-) the conservative threshold of PCL >= 33. Next, linear regression models assessed the effect of group on circadian IS and HR measures, using the control group as the reference group. Models accounted for the effect of sex, age, number of days with data, weeks since the event and minutes labelled as “sleep” as covariates. The regression models provided the p-values for comparing PTSD+ vs. control, and PTSD– vs. control, while Tukey’s HSD-adjusted p-values from a post-hoc test were used for comparing PTSD+ vs. PTSD–. For HR measures, six models were used to examine group differences in mean and standard deviation of MESOR, acrophase and amplitude, with significance determined by comparing each model to covariates-only model and using Bonfferoni correction for multiple comparisons (corrected α = 0.05/6 = 0.008). Linear regression models were also used to examine the relationship between variability in IS and HR measures and PCL-5 symptom severity at follow-up. To quantify the contribution of circadian measures to PTSD prediction above and beyond initial PTSD severity, PCL-5 scores from the first assessment time point were first regressed on follow-up PCL-5 scores and the residual PCL-5 was used as a dependent variable. Finally, a three-step hierarchical logistic regression analysis was conducted in order to assess the incremental contribution of IS and HR measures to future PTSD risk among survivors. The first regression step included sex, age, number of days with data, weeks since the event and minutes labelled as “sleep” as predictors. The second step incorporated IS as an additional predictor, and then HR MESOR mean was added as a predictor in the third step. Models were compared using analysis of deviance, and Tjur’s R^2^ value was used as the metric for pseudo R^2^.

## Results

### Trauma exposure and PTSD status

Out of 211 trauma survivors, data on trauma exposure was provided by 207 individuals. From this cohort, 195 participants (94.2%) reported being directly exposed to an immediate threat to their lives during the attack at the music festival. 158 participants (76.3%) also reported witnessing deceased or injured individuals during the traumatic event, and 136 participants (65.9%) indicated that they witnessed injuries to their friends and loved ones. 148 participants (71.5%) reported on the loss of loved ones during the event. The notably high prevalence of extreme trauma exposure led to high PTSD rates at follow-up, with 97 survivors (54.2%) exceeding the conservative threshold of PCL >= 33 nine-to-eleven-month following the attack, hence labeled PTSD+. For the remining 82 survivors (45.8%), PCL-5 scores at follow-up were lower than 33, hence labeled PTSD-. For group comparisons across time points see **Table 1**. For analysis of additional covariates see **Supplementary Table S1**.

### Circadian sleep instability and PTSD status

Sleep interday stability (IS) scores were positively skewed with values ranging between 0.21 to 0.82 and a mean (SD) of 0.57 (0.12). Out of a total of 282 participants with IS scores, for 216 participants (76.5%) IS scores were above 0.5, indicating that for most participants the variance within each day was lower compared to their overall variance during the whole measurement period (i.e., consistent rhythmicity in sleep-wake cycles). Participants slept on average seven hours and sixteen minutes (30.2% of total number of minutes per day) with a standard deviation of 43 minutes per day. For more information see **Supplementary material**. Multiple linear regression model yielded a significant effect of group on IS (F(_2, 251_) = 28.72, p < .0001), adjusted for sex, age, number of days of data, weeks since the event and mean minutes labelled as “sleep”. This effect was driven by lower IS scores in survivors with PTSD at follow-up compared to survivors with no PTSD (B = -0.05, 95% CI -0.1 to - 0.09, t_(251)_ = -2.96, p = .009), as well as compared to controls (B = - 0.14., 95% CI -.018 to -0.09, β = -1.1, t_(251)_ = -5.9, p <.001) (**Figure 1B**). Survivors with no PTSD also exhibited lower IS compared to control (B = -0.08, 95% CI -.013 to - 0.04, β = -0.66, t_(251)_ = -3.4, p = .001). The model also uncovered significant effects for age and weeks since the event (B = -0.002, 95% CI -.0002 to -.0045, β = -0.13, t_(251)_ = -2.2, p = .029; B = -0.002, 95% CI -.003 to -.000002, β = -0.16, t_(251)_ = -1.9, p = .050, respectively), but not for sex, number of days of data, and minutes labelled as “sleep” (all p’s > .216).

Next, a multiple linear regression model examined whether IS is associated with PTSD symptom severity among survivors at follow-up, while adjusting for same covariates used in the previous section. As depicted in **Figure 1C**, the effect of IS was significant (B = -28.94, 95% CI -49.28 to – -8.60, β = -0.21, t_(154)_ = -2.81, p = .006), indicating that sleep instability was associated with higher future PTSD symptom severity. Finally, using PCL-5 residual score at follow-up as a dependent variable after accounting for PCL-5 scores from the first assessment also yielded a significant prediction model (B = -16.14, 95% CI -29.20 to -3.07, β = -0.19, t_(156)_ = -2.44, p = .016). Hence, IS contributed to the prediction of future PTSD symptom severity above and beyond initial PTSD severity. See **Supplementary Table S2&S3** for a full description of the regression models.

### Circadian HR instability and PTSD

Across the whole sample, mean (SD) MESOR was 75.01 (7.75) BPM. On average, HR peaked on average at 17:53 PM (SD: 16:13-19:33), with a mean (SD) increase of 10.77 (2.64) BPM, representing phase and amplitude, respectively. For variable distribution and full correlation matrix see **Supplementary Figures S2&S3**. To examine the effect of group on circadian rhythm parameters, six separate multiple regression models were performed with circadian parameters as the dependent variables and group and the above covariates as independent predictors. Model selection resulted in two models in which group effect was significant. Specifically, group effects were significant in models predicting mean MESOR (p <.001) and acrophase SD (p <.001) (see **Supplementary Table S4** for model selection parameters). For MESOR mean, the effect was driven by higher MESOR values in survivors with PTSD at follow-up (M = 78.6, SD = 7.2) compared to survivors with no PTSD (M = 73.7, SD = 6.91; B = 3.58, 95% CI 0.83 to 6.33, t_(251)_ = 3.07, p = .007), as well as compared to controls (M = 72.6, SD =8.0; B = 5.13, 95% CI 2.3 to 8.0, β = 0.65, t_(251)_ = 3.5, p <.001) (**Figure 2B**). The difference between survivors with no PTSD and controls was not significant (B = 1.5, 95% CI --1.42 to 4.54, β = 0.2, t_(251)_ = 1.0, p = .305). The effect for acrophase SD (i.e., stability in HR peak time) was driven by higher circadian HR instability in survivors with PTSD (M = 0.83 , SD = 0.37) as well as in survivors with no PTSD (M = 0.76, SD = 0.32), compared to controls (M = 0.61, SD = 0.34; B = 0.23, 95% CI 0.09 to 0.37, β = 0.66, t_(251)_ = 3.29, p =.001; B = 0.19, 95% CI 0.05 to 0.34, β = 0.54, t_(251)_ = 2.58, p = .010, respectively). Survivors with PTSD were not statistically different from survivors with no PTSD in acrophase SD (p = .727) (**Figure 2C**). No other covariates were significant in the selected models. For a full description of all models see **Supplementary Tables S5**&**S6**.

Next, a multiple regression model examined whether MESOR mean and acrophase SD are associated with PTSD symptom severity among survivors at follow-up, while adjusting for same covariates used in the previous section. Both models yielded significant effects (F_(6,152)_ = 5.62, p = <.001; F_(6,152)_ = 4.81, p <.001, respectively). Examining model coefficients revealed that MESOR mean was a significant predictor for future PTSD among survivors (B = 0.45, 95% CI 0.09 to 0.82, β = 0.20, t_(152)_ = 2.47, p = .017) while acrophase SD was not (p = .161). In the latter, model significance was mainly due to a significant sex effect (p = .002). Using residual PTSD scores after accounting for PCL-5 scores from the first assessment eliminated the effect of MESOR mean on PTSD severity (p = .324). For a full description of the severity model see **Supplementary Table S7**.

### PTSD risk prediction

Finally, a hierarchical logistic regression analysis showed that in a model that includes sex, age, number of days with data, weeks since the event and minutes labelled as “sleep” as covariates, sex (OR = 0.40, 95% CI 0.2 to 0.78, p = .008) and number of days (OR = 0.94, 95% CI 0.90 to 0.99, p = .024) significantly predict future PTSD risk (i.e., PTSD group classification at follow-up) (Tjur’s R² = 0.13). Importantly, adding IS to the model significantly improved its fit (χ²_(1)_ = 5.87, p = .016, Tjur’s R² = 0.16), with IS significantly predicting PTSD status at follow-up (OR = 0.03, 95% CI 0.00 to 0.50, p = .019). Finally, adding MESOR mean further improved the fit of the model, achieving the highest predictive value (χ²_(1)_ = 6.33, p = .011, Tjur’s R² = 0.20), with MESOR mean emerging as a significant predictor (OR = 1.07, 95% CI 1.02 to 1.13, p = .015). For a full description of model coefficients see **Supplementary Table S8.**

## Discussion

The current study focused on behavioral and physiological indices of circadian (in)stability that were extracted from a full month recording in real-life settings in a large sample of young adults, three to six months following their exposure to mass trauma. Findings suggest that, compared to a matched control group, survivors of mass trauma exhibit reduced sleep interday stability (IS). In other words, they exhibit instability in their daily circadian sleep-wake cycles. Moreover, within the survivors’ group lower IS scores predicted increased likelihood to develop PTSD and more severe PTSD symptoms eight to eleven months following trauma exposure. Elevated physiological circadian instability in the form of higher standard deviation in heart rate peak time (acrophase) was also found among trauma survivors compared to controls, as well as elevated basic heart rate (MESOR), which was more severe is survivors with PTSD. Together, these behavioral and physiological indices of circadian disruptions incrementally and uniquely contributed to the prediction of future PTSD risk among survivors. These findings provide compelling evidence that circadian instability in the aftermath of mass trauma exposure predicts elevated future PTSD risk.

The unique cohort of the current study includes survivors of the tragic surprise terror attack on the Supernova music festival on October 7th, 2023. During the attack, hundreds of civilian festival attendances were murdered, with many others wounded and kidnaped. The survivors were subjected to extreme trauma that involved a real and imminent danger to their lives and the lives of festival attendances around them. Following this horrifying event, 63.7% of the survivors in our cohort exhibited PTSD symptom severity at a level above the clinical cutoff for PTSD, three to six months following the event. Eight to eleven months post event 54.2% of the survivors still exhibited symptom severity above the clinical cutoff for PTSD. Such substantial rate of PTSD is higher than the previously reported ∼40% PTSD rate in the first six months following terror-attacks such as the 9/11 attack on the New York Trade Center (2, 36). These elevated levels of trauma symptoms among survivors may reflect the severity and nature of the traumatic event as well as the fact that its aftermath included the onset of a war in Israel. Indeed, in a representative sample of the general population in Israel, PTSD symptom severity level was doubled from before to after the October 7^th^ terror attack (37). Hence, the painful continuation of the war could account for the slow rate of recovery among survivors in our cohort.

Within this traumatized cohort, a full month of wearable recording in real-life settings, three-to-six months post-event, enabled prediction of PTSD status and symptom severity eight-to-eleven months post-event. Specifically, lower consistency (i.e., instability) in sleep-wake cycles in the aftermath of trauma predicted higher likelihood for future development of PTSD and more severe PTSD symptom severity. Critically, this prediction remained meaningful even after controlling for initial levels of PTSD symptoms. This finding extends previous studies demonstrating that sleep disturbances at close proximity to trauma exposure, immediately prior to or following it, predict increased likelihood for subsequent development of PTSD (11, 13, 15–17, 21, 38). Interestingly, the majority of early work relied on self-report measures of sleep disturbances, while studies implementing objective measurements often failed to distinguish PTSD from healthy controls. Here, by using a long measurement period of a full month recording in natural settings and quantifying sleep disturbances as a function of circadian instability rather than mean sleep scores, we were able to uncover a strong positive association between objective measures of sleep instability and future PTSD status and symptom severity. These findings align with the notion of posttraumatic chronodisruption (11), highlighting sleep interday instability as a robust indicator of trauma-related circadian disruption. Interestingly, interday stability was found to differentiate insomnia patients from chronic PTSD patients, suggesting that it may reflect trauma specific index of circadian disruption (21). One potential trauma related feature that may substantially contribute to circadian instability is nightmares, given their unpredictable frequency and occurrences (39).

In addition to behavioral circadian instability, current results also highlight the role of physiological measures of circadian rhythm and their stability in predicting future PTSD risk. Specifically, both higher basic levels of HR (MESOR) and higher variability of HR peak time (acrophase) at trauma aftermath were associated with more severe PTSD symptoms at follow-up. These findings align with and extend vast literature on the role of autonomic dysregulation in PTSD pathophysiology (40, 41). For example, measures of heart rate variability (HRV) during the 8-week post traumatic event emerged as important features in predicting PTSD status (42). Critically, here higher MESOR uniquely contributed to PTSD risk above the contribution of sleep-wake cycle instability. Previous research suggested that the exact mechanism linking sleep disorders and autonomic dysregulation can involve a shared factor or that there may be a causal link where sleep leads to autonomic disfunction (43). Regardless of their causal relations, our results indicate that the presence of both circadian autonomic dysregulation and circadian behavioral sleep instability in the aftermath of trauma is associated with incremental risk for future PTSD. These findings underscore the importance of using multiple domains in understanding the pathophysiology of PTSD, particularly with respect to posttraumatic chronodisruption.

While providing vital insights, this study has several limitations. First, the use of consumer wearables limits our ability to infer on sleep stages. This is important given extensive evidence links REM sleep with PTSD (44). Second, the fact that participants had access to their data might have influenced the results. Although all notifications were turned off and participants were requested to avoid interacting with the sensor, it is still possible that participants were aware of their measures. Third, the observational nature of this study cannot account for causality. To this end, it is possible that some survivors had sleep disturbances already prior to their trauma exposure, which in turn might have increased their vulnerability for PTSD.

In conclusion, this study highlights the significant role of circadian instability, particularly in sleep and heart rate, in predicting elevated risk for future PTSD and more severe symptoms among survivors of mass trauma. Our findings emphasize the importance of maintaining regular circadian rhythms, both behaviorally and physiologically, to mitigate the adverse psychological impacts of traumatic events. Future longitudinal studies are essential to further elucidate the complex interplay between autonomic dysregulation, sleep patterns, and PTSD symptoms. These insights, in turn, could contribute to the development of targeted interventions that leverage wearable technology for monitoring and improving mental health outcomes in trauma-exposed populations.

## Biographical note

### Data availability statement

The data that support the findings of this study are available from the corresponding author R.A. upon reasonable request.

## Acknowledgments

The writers desire to express gratitude to the numerous individuals who facilitated this endeavor during challenging times in Israel. The SafeHeart NGO administration and initial volunteers (Karina Dessau, Reut Plonsker, Tal Zagursky, Yair Grynbaum, Guy Simon, Nir Tadmor, Igal Tartakovsky, Irit Hacmun, Stephanie Cohen, Shiran Maor, Dr. Demian Halperin & Efrat Atun) who labored diligently to ensure care for the beneficiaries. We appreciate all the SafeHeart volunteers, healthcare professionals, and the investigative team. Lastly, we aim to convey our profound appreciation to the remarkable survivors of the event who bestowed upon us their confidence and exerted a monumental endeavor to assist others even amidst such arduous circumstances.

