## Supplementary Material for "Circadian instability predicts PTSD symptom severity following mass trauma"

Supplemental Methods

*Sensor data preprocessing:* Sleep data preprocessing involved the identification of sleep and wake periods as provided by the sensor. Periods where sleep data could not be determined were also identified. Specifically, if the period between the wake-up time and the next sleep onset time exceeded 30 hours, making it impossible to detect sleep in this given day, all the minutes in this epoch were labeled as "unknown". The result was a 5-minute resolution vector in which each minute was labeled as "sleep," "awake," or "unknown". For sleep stability analysis, days with unknowns were omitted. Furthermore, following (1), participants with less than seven full days of available data were also excluded.

HR data preprocessing involved omitting subjects that were omitted in the sleep stability step. HR data was then down-sampled to a 5-minute resolution. Since the analyses focused on HR circadian components, only full data days were included (ranging from 12:00 pm to 11:59 am). Several steps were taken to balance the need to exclude days with excessive missing data while maximizing the number of days included per participant. First, periods of missing data were identified, and days with more than 37% (540 minutes) of missing data were discarded. Data on physical activity (steps per minute) contained a notable amount of missing data. Since steps were included as a covariate in the circadian model, minutes that contained HR samples but lacked steps data were assigned with zero steps.

*Circadian sleep instability assessment:* Sleep inter-day stability (IS) was assessed using the following formula (1):

$$IS=\frac{N\sum_{m=1}^{p} \left( \bar{X}_{m}-\bar{X} \right)^{2}}{p\sum_{i=1}^{N} \left( \bar{X}_{i}-\bar{X} \right)^{2}}$$

where N is the total number of data points; p is the number of data points per day (here 5-m epochs = 288 data points per day); $\overline{X}$ is the overall mean; $\overline{X}_{m}$ is the epoch-mean across all days; and X_i_ represents the individual minute.

To examine the robustness of IS measure, the effect of data resolution (1 minutes, 5 minutes and 60 minutes) was examined as well as IS sensitivity to the number of available days with data (full recording duration, first 7 days, first 15 days). Resolution had nearly no effect on IS ranking but did have a small effect on overall IS score, with correlation equal to r = 0.99 between different resolutions and a resolution of 60 minutes resulting in the highest group mean IS scores (M = 0.6, SD = 0.14). Changing the number of days used for IS calculation also had a small effect on total IS scores and ranking, with high correlations between whole sample IS and the first 15 days or the first 7 days (r = 0.94, p <.001; r = 0.82, p <.001, respectively). To maximize data availability and avoid overestimation of sleep stability, the final IS set was based on the full recording duration and a 5-minute resolution.

Correlation analyses tested putative associations between individual IS scores and additional sleep measures including average and standard deviation of sleep duration and sleep onset. As expected, IS scores were correlated with sleep standard deviation measures, but not with mean sleep measures. See **Figure S1** for a full correlation matrix.

*Circadian HR assessment:* The cosinor model was used for assessing HR circadian rhythms (2):

$$Y \left( t \right)=M+K \cos\left( \frac{2\pi t}{\tau}+\varphi\right)+ d+e$$

M is known as the MESOR (intercept), K is the amplitude and φ is the acrophase (phase) of the circadian rhythm. τ is the period (288 points in our case), d is the linear regressor of activity.

Across the six cosinor circadian parameters (mean and standard deviation of MESOR, amplitude and acrophase), correlation was highest between standard deviations of MESOR and of amplitude (r = -0.59, p <.001). HR parameters were only weakly correlated to the number of available days and average missing data segment (maximum r = -0.21, minimum p =.001). See **Figure S2** for distribution of cosinor parameters.

Supplemental Results

*Contribution of additional trauma-related variables:* To assess the contribution of other well-known risk and resilience factors for future PTSD risk, group differences were examined in relation to the experiencing of additional crises and the participation in any form of psychotherapy since the attack to follow-up assessment, the experiencing of adverse childhood experiences (3), and physical state and drug intake during the one month sensor recording period. With respect to additional crisis, at follow-up, 79 participants (44.1%) reported experiencing additional crisis after the October 7^th^ attack. Out of these, 27 participants reported experiencing personal life stressors (mainly divorce or breakup), 19 participants reported on death of closed ones, of which 8 reported of war-related death. Additional 8 participants reported of being in accidents or participating in violent events, 8 reported financial and occupational difficulties, 6 reported on war-related events (e.g. being recruited to reserve duty or family being exposed to direct missile attack), 4 reported on serious illness of closed one, 2 reported on experiencing psychotic episodes, one participant reported of witnessing others killed while serving in the reserve force and 15 participants did not specify the type of crisis they experienced. Survivors with PTSD at follow-up reported experiencing additional crises after October 7^th^ attack at a higher prevalence (54.6%) than survivors without PTSD (31.7%; χ²_(1)_ = 8.57, p = .003). With respect to participating in psychotherapy, almost all participants reported receiving psychotherapy since the event, but survivors with PTSD indicated that the treatment was less helpful (M = 59.07; SD = 29.3) compared to survivors without PTSD (M =77.6; SD = 22.4; t_(165.64)_ = 4.66, p < .001). The groups did not differ in terms of exposure to adverse childhood experiences (ACE), and data on physical state and regular drug intake was insufficient for statistical comparisons. See **Table S1** for full results.

Supplemental Figures

**Figure S1: Sleep inter-day stability and sleep related factors**

**Figure S1**. **Correlation matrix between sleep inter-day stability (IS) and traditional sleep measures.** *IS =* inter-day stability*; days =* number of days used for IS calculations*; duration_m =* mean main sleep duration (minutes)*; onset_m =* mean sleep onset (minutes since midnight)*; average_bout =* average sleep bout*; MeanMinutesAsSleep =* Mean minutes labelled as “sleep”*; onset_sd =* standard deviation of sleep onset (minutes since midnight)*; duration_sd =* standard deviation of main sleep duration*;* main sleep onset and duration were defined as the longest night sleep which started between 8pm and 8am and lasted at least three hours. Shown here are unadjusted p-values. Inignificant = p >.05.

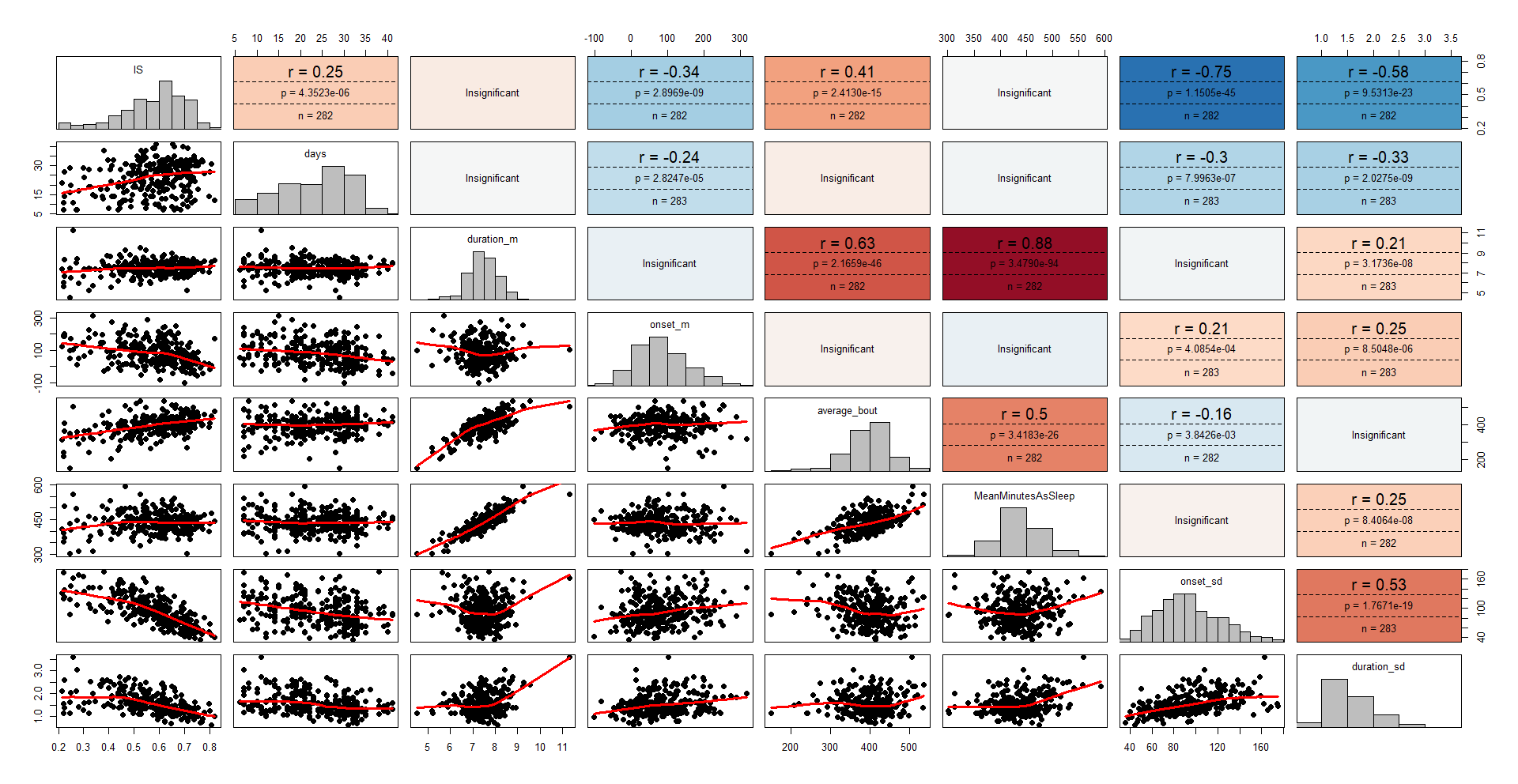

**Figure S2: Cosinor variables distribution**

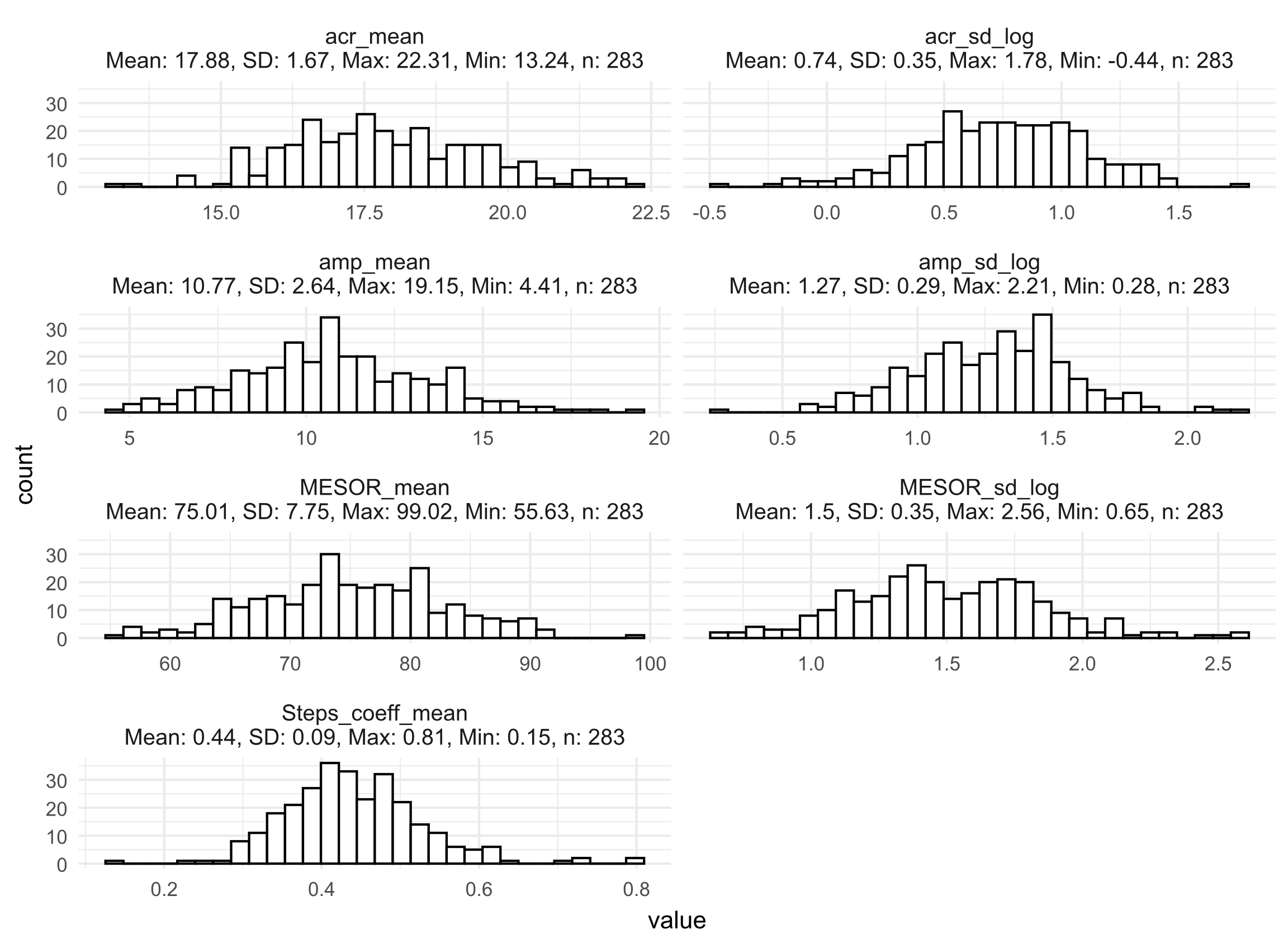

**Figure S2**. **Circadian feature distribution.** *Acr =* acrophase (heart rate peak time, in fractional hours)*, amp =* heart rate amplitude; *MESOR =* midline estimation of rhythm; *Steps_coeff =* coefficient of the steps count covariate, included in the model; *_sd_log =* standard deviation, log transformed. For acrophase, circular mean and standard deviations were used.

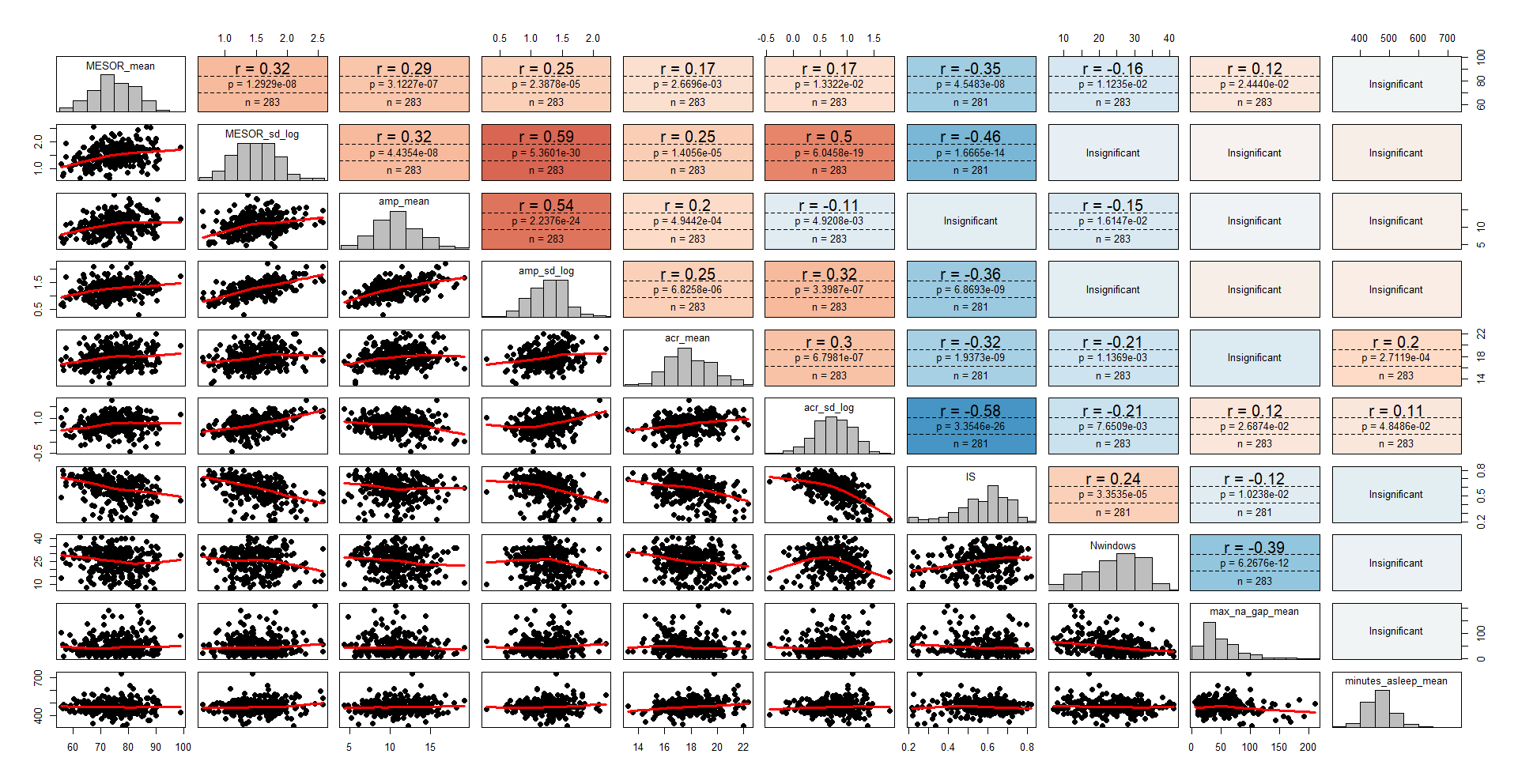
**Figure S3: Associations between heart rate circadian features, sleep inter-day stability and data availability**

**Figure S3. Correlation matrix between heart rate circadian features, sleep inter-day stability (IS) and data availability.** *acr =* acrophase (heart rate peak time, in fractional hours)*, amp =* heart rate amplitude; *MESOR =* midline estimation of rhythm; *_sd_log =* standard deviation, log transformed. For acrophase, circular mean and standard deviations were used. *IS =* inter-day (sleep) stability; *Nwindows* = number of windows (days) used for physiological features calculation; *max_na_gap_mean* = mean of the longest period of missing data, in minutes; *minutes_asleep_mean* = mean minutes labelled as “sleep”. Shown here are unadjusted p-values. Insignificant = p >.05.

Supplemental Tables

| **Table S1: Additional trauma-related variables** | | | | | |
| --- | --- | --- | --- | --- | --- |
|  | Survivors  PTSD + | Survivors  PTSD - | Control | Survivors Dropout | p-value |
| Total n | 97 | 82 | 113 | 33 |  |
| ACE, median (IQR) | 2 (2.5) | 1 (3) | 2 (2.5) | 1 (2) | .068 |
| **During the assessment** |  |  |  |  |  |
| Physical state during sensor recordings |  |  |  |  |  |
| Wounded, n (%) | 2 (2.06%) | 0 (0%) | 1 (0.88%) | 1 (3.03%) | - |
| In pain, n (%) | 14 (14.43%) | 3 (3.65%)) | 11 (9.73%) | 4 (12.12%) | - |
| mobility-impaired, n (%) | 5 (5.15%) | 5 (6.09%) | 3 (2.65%) | 6 (18.18%) | - |
| **Follow up assessment** | | | | | |
| Therapy since event, n (%) | 94 (96.9%) | 78 (95.1%) | - | - | - |
| Therapy help (1-100), mean (sd) | 59.07 (29.3) | 77.6 (22.4) | - | - | **<.001** |
| Another crisis since October 7^th^, n (%) | 54 (55.7%) | 27 (32.9%) | - | - | **.003** |
| ^1^ p-values are based on chi square test for categorical variables, Kruskal–Wallis tests for ACE scores, and student’s t-test for two groups comparisons.  ^2^ number of participants per variable ranged between 179 -300  ^3^ The percentage in parentheses is taken from the total available data for this variable.  ^4^ physical state was not statistically compared due to insufficient N per cell  ^5^ Abbreviations: *ACE =* adverse childhood experience (3) questionnaire, *IQR* = interquartile range;  ^6^ prior trauma = *“Have you experienced a traumatic event in the past, other than the October 7 attack?”.* participants who chose not to responded were distributed between groups in the following way: Survivors PTSD +, n = 4; Survivors PTSD -, n = 4; Survivors Dropout, n = 0; Control, n= 1.  Therapy since = “*Have you received psychological or mental health treatment since October 7?”*  Therapy help = “*To what extent do you feel that the treatment has helped you so far?”*  Another crisis since October 7^th^ = *“Have you experienced another crisis after the events of October 7? For example – a breakup/divorce, the loss of a loved one, job loss, a car accident, an injury, a violent incident, or similar.”.* | | | | | |

| **Table S2: Group differences in sleep inter-day stability** | | | | | | |
| --- | --- | --- | --- | --- | --- | --- |
| *Predictors* | *B* | *SE* | *β* | *CI* | *t* | *p* |
| (Intercept) | .57 | .09 | .57 | .39 – 0.75 | 6.3 | **<.001** |
| Group: PTSD - | -.08 | .02 | -.66 | -.13 – -.03 | -3.4 | **.001** |
| Group: PTSD + | -.14 | .02 | -1.1 | -.18 – -.09 | -5.9 | **<.001** |
| Sex: male | -.00 | .01 | -.04 | -.03 – .02 | -0.3 | .781 |
| Age | .00 | .00 | .13 | .00 – .00 | 2.2 | **.029** |
| Data availability (days) | .00 | .00 | .08 | -.00 – .00 | 1.2 | .216 |
| Mean minutes labelled as “sleep” | .00 | .00 | .02 | -.00 – .00 | 0.3 | .743 |
| weeks since event | -.00 | .00 | -.16 | -.00 – -.00 | -1.9 | **.050** |
| Observations | 259 | | | | | |
| R^2^ / R^2^ adjusted | 0.215 / 0.193 | | | | | |
| *Note.* Results of linear regression model predicting sleep inter-day stability (IS). Model significance: F(7,251) = 9.81, p = <.001. *PTSD -/+* : participants were assigned to PTSD groups based on their follow-up PCL-5 scores, using a cutoff of 33. Selected reference group - control. The remaining comparison (PTSD + vs PTSD -) was performed using a post-hoc test and the p-value was adjusted using Tukey’s HSD. *SE =* standard error. *CI =* 95% confidence interval. ‘Weeks since event’ refers to the time from October 7^th^ and the start of sensor measuring period. | | | | | | |

| **Table S3: Sleep inter-day stability and PTSD symptom severity** | | | | | | |
| --- | --- | --- | --- | --- | --- | --- |
| *Predictors* | *B* | *SE* | *β* | *CI* | *t* | *p* |
| (Intercept) | 29.75 | 18.80 | .23 | -7.39 – 66.90 | 1.58 | .116 |
| Sleep inter-day stability (IS) | -28.94 | 10.30 | -.21 | -49.28 – -8.60 | -2.81 | **.006** |
| Sex: male | -8.04 | 2.53 | -.48 | -13.04 – -3.03 | -3.17 | **.002** |
| Age | -.31 | .21 | -.12 | -.72 – .10 | -1.51 | .132 |
| Data availability (days) | .04 | .03 | .11 | -.01 – .09 | 1.46 | .146 |
| Mean minutes labelled as “sleep” | .42 | .27 | .12 | -.11 – .96 | 1.56 | .121 |
| weeks since event | 29.75 | 18.80 | .23 | -7.39 – 66.90 | 1.58 | .116 |
| Observations | 159 | | | | | |
| R^2^ / R^2^ adjusted | 0.167 / 0.140 | | | | | |
| *Note.* Results of linear regression model predicting PTSD (PCL-5) symptom severity at follow-up for the survivors group. Model significance: F(5,153) = 6.51, p < .001. *SE =* standard error. *CI =* 95% confidence interval. ‘Weeks since event’ refers to the time from October 7^th^ and the start of sensor measuring period. | | | | | | |

| **Table S4: Circadian HR model selection** | | |
| --- | --- | --- |
| Dependent variable | F(2,251) | p |
| MESOR (mean) | 8.14 | **<.001** |
| Amplitude (mean) | 0.79 | . 453 |
| Acrophase (mean) | 1.76 | .173 |
| MESOR (SD) | 3.76 | .024 |
| Amplitude (SD) | 0.37 | .686 |
| Acrophase (SD) | 5.51 | **.004** |
| *Note.* Result of hierarchical regression F-test comparing covariates-only model to model including PTSD group. Model is selected if it showed a significant effect at α = 0.05/6 = 0.008. | | |

| **Table S5: Selected regression models for group differences in circadian HR measures** | | | | | | | | | | | | |
| --- | --- | --- | --- | --- | --- | --- | --- | --- | --- | --- | --- | --- |
|  | **Model 1: MESOR mean**  F(7,251) = 10.82, p < .001 | | | | | | **Model 2: Acrophase SD (log)**  F(7,251) = 3.65, p < .001 | | | | | |
| *Predictors* | *B* | *SE* | *β* | *CI* | *t* | *p* | *B* | *SE* | *β* | *CI* | *t* | *p* |
| (Intercept) | 82.4 | 5.43 | .04 | 71.8 – 93.1 | 15.2 | **<.001** | 0.3 | .27 | -.28 | -0.21 – 0.83 | 1.2 | .243 |
| Group: PTSD - | 1.5 | 1.51 | .20 | -1.42 – 4.54 | 1.0 | .305 | 0.2 | .07 | .54 | 0.05 – 0.34 | 2.6 | **.010** |
| Group: PTSD + | 5.1 | 1.46 | .65 | 2.3 – 8.00 | 3.5 | **<.001** | 0.2 | .07 | .66 | 0.09 – 0.37 | 3.3 | **.001** |
| Sex: male | -5.4 | .89 | -.68 | -7.12 – -3.6 | -6.0 | **<.001** | -0.0 | .04 | -.21 | -0.16 – 0.01 | -1.7 | .090 |
| Age | -0.1 | .07 | -.06 | -0.21 – 0.06 | -1.1 | .282 | 0.0 | .00 | .02 | -0.01 – 0.01 | 0.3 | .759 |
| Data availability (days) | -0.0 | .06 | -.04 | -0.16 – 0.09 | -0.6 | .556 | -0.0 | .00 | -.03 | -0.01 – 0.00 | -0.5 | .643 |
| Mean minutes labelled as “sleep” | -0.0 | .01 | -.05 | -0.03 – 0.01 | -0.9 | .379 | 0.0 | .00 | .08 | -0.00 – 0.00 | 1.3 | .200 |
| weeks since event | -0.0 | .06 | -.03 | -0.13 – 0.10 | -0.3 | .754 | 0.0 | .00 | .07 | -0.00 – 0.01 | 0.81 | .418 |
| Observations | 259 | | | | | | 259 | | | | | |
| R^2^ / R^2^ adjusted | 0.232 / 0.210 | | | | | | 0.092 / 0.067 | | | | | |
| *Note.* Results of the selected linear regression models predicting physiological circadian features. Participants were assigned to PTSD groups based on follow-up PCL-5 scores, using a cutoff of 33. Selected reference group - control. The remaining comparison (PTSD + vs PTSD -) was performed using a post-hoc test and the p-value was adjusted using Tukey’s HSD. *MESOR =* midline estimation of rhythm, *Acrophase SD:* circular standard deviation of heart rate peak time (log transformed); *Acrophase Mean:* circular mean of heart rate peak time; *SE =* standard error. *CI =* 95% confidence interval. ‘Weeks since event’ refers to the time from October 7^th^ and the start of sensor measuring period. | | | | | | | | | | | | |

| **Table S6: Unselected regression models for group differences in circadian HR measures** | | | | | | |
| --- | --- | --- | --- | --- | --- | --- |
|  | **Model 3: MESOR SD (log)**  F(7,251) = 2.49, p = .017 | | | | | |
| *Predictors* | *B* | *SE* | *β* | *CI* | *t* | *p* |
| (Intercept) | 1.4 | .27 | -.13 | 0.86 – 1.91 | 5.2 | **<.001** |
| Group: PTSD - | .09 | .07 | .24 | -0.06 – 0.23 | 1.1 | .250 |
| Group: PTSD + | .18 | .07 | .53 | 0.04 – 0.33 | 2.6 | **.010** |
| Sex: male | -.09 | .04 | -.25 | -0.17 – -0.0 | -1.2 | **.049** |
| Age | -.00 | .00 | -.09 | -0.01 – 0.00 | -1.4 | .165 |
| Data availability (days) | .00 | .00 | .03 | -0.00 – 0.01 | 0.4 | .665 |
| Mean minutes labelled as “sleep” | 0.00 | .00 | .02 | -0.00 – 0.00 | 0.3 | .733 |
| weeks since event | 0.00 | .00 | .13 | -0.00 – 0.01 | 1.5 | .142 |
| R^2^ / R^2^ adjusted | 0.065 / 0.039 | | | | | |
|  | **Model 4: mean amplitude**  F(7,251) = 6.75, p <.001 | | | | | |
| *Predictors* | *B* | *SE* | *β* | *CI* | *t* | *p* |
| (Intercept) | 12.8 | 1.89 | -.05 | 9.09 – 16.53 | 6.8 | **<.001** |
| Group: PTSD - | -.66 | .53 | -.25 | -1.70 – 0.37 | -1.3 | .209 |
| Group: PTSD + | -.45 | .51 | -.17 | -1.45 – 0.55 | -0.9 | .376 |
| Sex: male | 1.0 | .31 | .39 | 0.42 – 1.64 | 3.3 | **.001** |
| Age | -.13 | .02 | -.33 | -0.18 – -0.08 | -5.4 | **<.001** |
| Data availability (days) | -.02 | .02 | -.05 | -0.06 – 0.03 | -0.7 | .458 |
| Mean minutes labelled as “sleep” | .00 | .00 | .07 | -0.00 – 0.01 | 1.2 | .236 |
| weeks since event | .00 | .02 | .01 | -0.04 – 0.04 | 0.1 | .951 |
| R^2^ / R^2^ adjusted | 0.159 / 0.135 | | | | | |

| **Table S6 – Cont.** | | | | | | |
| --- | --- | --- | --- | --- | --- | --- |
|  | **Model 5: Amplitude SD (log)**  F(7,251) = 2.58, p = .013 | | | | | |
| *Predictors* | *B* | *SE* | *β* | *CI* | *t* | *p* |
| (Intercept) | 1.3 | 0.22 | .01 | 0.85 – 1.72 | 5.8 | **<.001** |
| Group: PTSD - | -.04 | 0.06 | -.13 | -0.16 – 0.08 | -0.6 | .539 |
| Group: PTSD + | .00 | 0.06 | .00 | -0.12 – 0.12 | 0.02 | .987 |
| Sex: male | .02 | 0.04 | .05 | -0.06 – 0.09 | .42 | .675 |
| Age | -.01 | 0.00 | -.22 | -0.02 – -0.00 | -3.4 | **.001** |
| Data availability (days) | .00 | 0.00 | .01 | -0.00 – 0.01 | 0.09 | .928 |
| Mean minutes labelled as “sleep” | .00 | 0.00 | .07 | -0.00 – 0.00 | 1.13 | .261 |
| weeks since event | .00 | 0.00 | .08 | -0.00 – 0.01 | 0.87 | .387 |
| R^2^ / R^2^ adjusted | 0.067 / 0.041 | | | | | |
|  | **Model 6: Acrophase mean**  F(7,251) = 4.85, p < .001 | | | | | |
| *Predictors* | *B* | *SE* | *β* | *CI* | *t* | *p* |
| (Intercept) | 15.9 | 1.23 | -.28 | 13.51 – 18.35 | 13.0 | **<.001** |
| Group: PTSD - | .47 | .34 | .29 | -0.20 – 1.15 | 1.4 | .168 |
| Group: PTSD + | .62 | .33 | .37 | -0.03 – 1.27 | 1.9 | .062 |
| Sex: male | .28 | .20 | .17 | -0.12 – 0.68 | 1.4 | .167 |
| Age | -.04 | .02 | -.17 | -0.07 – -0.01 | -2.7 | **.008** |
| Data availability (days) | -.01 | .01 | -.06 | -0.04 – 0.02 | -0.9 | .383 |
| Mean minutes labelled as “sleep” | .01 | .00 | .17 | 0.00 – 0.01 | 2.8 | **.006** |
| weeks since event | .01 | .01 | .06 | -0.02 – 0.03 | 0.6 | .521 |
| R^2^ / R^2^ adjusted | 0.119 / 0.095 | | | | | |
| Observations | 259 | | | | | |
| *Note.* Results of the unselected linear regression models predicting physiological circadian features. Participants were assigned to PTSD groups based on follow-up PCL-5 scores, using a cutoff of 33. Selected reference group - control. The remaining comparison (PTSD + vs PTSD -) was performed using a post-hoc test and the p-value was adjusted using Tukey’s HSD. *MESOR SD =* standard deviation of midline estimation of rhythm; *SD = standard deviation*; *SE =* standard error. *CI =* 95% confidence interval. ‘Weeks since event’ refers to the time from October 7^th^ and the start of sensor measuring period. All standard deviations variables were first log-transformed before entering the model. | | | | | | |

| **Table S7: HR circadian measures and PTSD symptom severity** | | | | | | |
| --- | --- | --- | --- | --- | --- | --- |
|  | **MESOR Model (dependent variable: PCL-5)** | | | | | |
| *Predictors* | *B* | *SE* | *β* | *CI* | *t* | *p* |
| (Intercept) | -4.9 | 23.07 | .16 | -50.50 – 40.67 | -0.2 | .832 |
| MESOR \ Acrophase SD (log) | .45 | .18 | .20 | 0.09 – 0.82 | 2.5 | **.015** |
| Sex: male | -5.4 | 2.74 | -.32 | -10.85 – -0.00 | -1.2 | **.050** |
| Age | -.29 | .21 | -.11 | -0.70 – 0.13 | -1.4 | .174 |
| Data availability (days) | -.31 | .18 | -.14 | -0.66 – 0.04 | -1.7 | .085 |
| Mean minutes labelled as “sleep” | .03 | .02 | .09 | -0.02 – 0.07 | 1.2 | .218 |
| weeks since event | .25 | .28 | .07 | -0.29 – 0.80 | 0.9 | .358 |
| R^2^ / R^2^ adjusted | 0.182 / 0.149 | | | | | |
|  | **Acrophase (SD) model (dependent variable: PCL-5)** | | | | | |
| *Predictors* | *B* | *SE* | *β* | *CI* | *t* | *p* |
| (Intercept) | 27.32 | 18.50 | .23 | -9.24 – 63.87 | 1.5 | .142 |
| MESOR \ Acrophase SD (log) | 5.15 | 3.65 | .11 | -2.07 – 12.36 | 1.4 | .161 |
| Sex: male | -7.99 | 2.54 | -.48 | -13.01 – -2.98 | -3.1 | **.002** |
| Age | -.36 | 0.21 | -.14 | -0.77 – 0.06 | -1.7 | .091 |
| Data availability (days) | -.30 | 0.18 | -.13 | -0.66 – 0.06 | -1.6 | .102 |
| Mean minutes labelled as “sleep” | .03 | 0.02 | .09 | -0.02 – 0.07 | 1.2 | .238 |
| weeks since event | .30 | 0.28 | .09 | -0.25 – 0.85 | 1.1 | .284 |
| R^2^ / R^2^ adjusted | 0.160 / 0.126 | | | | | |
| Observations | 159 | | | | | |
| *Note.* Results of linear regression models predicting PTSD (PCL-5) symptom severity at follow-up for the survivors group. Models’ significance: MESOR model: F(6,152) = 5.62, p = <.001 ; Acrophase (SD) model: F(6,152) = 4.81, p <.001. *MESOR =* midline estimation of rhythm, *Acrophase SD:* circular standard deviation of heart rate peak time (log transformed); *SE =* standard error. *CI =* 95% confidence interval. ‘Weeks since event’ refers to the time from October 7^th^ and the start of sensor measuring period. | | | | | | |

| **Table S8: PTSD risk models** | | | | |
| --- | --- | --- | --- | --- |
|  | **Model 1: covariates only** | | | |
| *Predictors* | *OR* | *CI* | χ² | *p* |
| (Intercept) | 4.04 | .09 – 177.2 | .73 | .467 |
| Sex: male | .40 | .20 – .78 | -2.7 | **.008** |
| Age | .96 | .91 – 1.02 | -1.3 | .208 |
| Data availability (days) | .94 | .90 – .99 | -2.3 | **.024** |
| Mean minutes labelled as “sleep” | 1.00 | 1.00 – 1.01 | .56 | .575 |
| weeks since event | 1.05 | .99 – 1.12 | 1.6 | .102 |
| R2 Tjur | 0.129 |  |  |  |
|  | **Model 2: covariates + IS** | | | |
| *Predictors* | *OR* | *CI* | χ² | *p* |
| (Intercept) | 23.6 | .37 – 1718.28 | 1.5 | .140 |
| Sex: male | .39 | .19 – .78 | -2.6 | **.008** |
| Age | .98 | .92 – 1.04 | -0.7 | .452 |
| Data availability (days) | .95 | .90 – 1.00 | -2.1 | **.039** |
| Mean minutes labelled as “sleep” | 1.00 | 1.00 – 1.01 | 0.5 | .598 |
| weeks since event | 1.04 | .98 – 1.11 | 1.2 | .237 |
| IS | .03 | .00 – .50 | -2.3 | **.019** |
| R2 Tjur | 0.163 |  |  |  |
|  | **Model 3: covariates + IS + MESOR mean** | | | |
| *Predictors* | *OR* | *CI* | χ² | *p* |
| (Intercept) | .07 | .00 – 35.38 | -0.8 | .406 |
| Sex: male | .56 | .26 – 1.19 | -1.5 | .129 |
| Age | .98 | .92 – 1.04 | -0.7 | .512 |
| Data availability (days) | .95 | .90 – 1.00 | -2.0 | **.042** |
| Mean minutes labelled as “sleep” | 1.00 | .99 – 1.01 | 0.4 | .655 |
| weeks since event | .04 | .98 – 1.11 | 1.2 | .210 |
| IS | .05 | .00 – 1.16 | -1.8 | .068 |
| MESOR mean | 1.07 | 1.02 – 1.13 | 2.4 | **.015** |
| R2 Tjur | 0.202 |  |  |  |
| Observations | 158 | | | |
| *Note.* Results of the hierarchical logistic regression models predicting PTSD status among survivors. Participants were assigned to PTSD groups based on follow-up PCL-5 scores, using a cutoff of 33. OR = Odds ratio; IS = inter-day stability; *MESOR Mean =* mean of midline estimation of rhythm. *SE =* standard error. *CI =* 95% confidence interval. ‘Weeks since event’ refers to the time from October 7^th^ and the start of sensor measuring period. | | | | |
